# Redefining kidney disease: Clinico-pathological and molecular findings from the Kidney Precision Medicine Project

**DOI:** 10.64898/2026.02.24.26347022

**Authors:** Christine P. Limonte, Jennifer A. Schaub, Robin Fallegger, Rajasree Menon, Insa M. Schmidt, Ian H. de Boer, Chirag Parikh, Charles E. Alpers, M. Luiza Caramori, Sylvia Rosas, Amy Mottl, Frank Brosius, Katherine Tuttle, James Lash, Julio Saez-Rodriguez, Laura H. Mariani, Ana C. Ricardo, Michael T. Eadon, Wenjun Ju, Joel Henderson, Laura Barisoni, Jeffrey B. Hodgin, Leila R. Zelnick, Kumar Sharma, Jeff Spraggins, Anand Srivastava, Sarah Schrauben, Matthew Weir, Chi-yuan Hsu, Tanika Kelly, Jonathan Taliercio, Hernan Rincon-Choles, Ruth Dubin, Debbie L. Cohen, Dawei Xie, Jing Chen, Jiang He, Amanda H. Anderson, Matthias Kretzler, Jonathan Himmelfarb, the CRIC Study Investigators, the Kidney Precision Medicine Project

## Abstract

**Background:** The Kidney Precision Medicine Project (KPMP) consortium aims to redefine chronic kidney disease (CKD) by integrating clinical, pathological, and molecular tissue data from kidney biopsies. Here, we demonstrate how biopsy data in CKD can clarify disease etiology and contribute to understandings of disease pathophysiology and clinical prognosis.

**Methods:** The KPMP is obtaining research kidney biopsies from individuals with CKD (defined as an estimated glomerular filtration rate [eGFR] < 60 mL/min/1.73m^2^ and/or albuminuria ≥30 mg/g creatinine) and diabetes (enrolled as diabetes and CKD or DKD) or hypertension (enrolled as hypertension and CKD or HCKD). A team of kidney pathologists and nephrologists adjudicated the primary clinico-pathological diagnosis for 258 participants with CKD. We compared pathological features and kidney transcriptional signatures between participants with a primary adjudicated diagnosis of diabetic nephropathy and those with other causes of CKD. We developed a model using clinical and biomarker data that predicted the probability of diabetic nephropathy and tested associations of the signature with CKD progression among Chronic Renal Insufficiency Cohort (CRIC) participants with diabetes (n=229).

**Results:** Among 183 participants enrolled as DKD, 102 (56%) had a primary adjudicated clinico-pathologic diagnosis of diabetic nephropathy. Among 75 participants enrolled as HCKD, 42 (56%) had a primary diagnosis of hypertension-associated kidney disease. Those with diabetic nephropathy, compared with other diagnoses, had more severe interstitial fibrosis, tubular atrophy, tubular injury, segmental sclerosis, and severe arteriolar hyalinosis, and single-nucleus and single-cell transcriptional analyses revealed upregulation of immune and inflammatory pathways and downregulation of oxidative phosphorylation. A combination of age, hemoglobin A1c, urine albumin-creatinine ratio, and serum KIM-1 and sTNFR1 predicted a clinico-pathologic diagnosis of diabetic nephropathy in the KPMP (AUC 0.82, 95% CI 0.75-0.89) and was associated with an increased risk of CKD progression among patients with diabetes enrolled in CRIC (HR 1.48 [95% CI 1.27-1.73] per 10% higher predicted probability of diabetic nephropathy).

**Conclusion:** In common presentations of CKD, kidney biopsies may alter a priori impressions, reveal a diversity of diagnosis, structure, and function that is associated with clinical outcomes and can impact therapeutic decisions.

## INTRODUCTION

Chronic kidney disease (CKD), defined as an estimated glomerular filtration rate (eGFR) <60 mL/min/1.73m^2^ and/or a urine albumin creatinine ratio (UACR) ≥30 mg/g, is associated with high morbidity and mortality, particularly for people who progress to kidney failure.^1^ Diabetes and hypertension are the most common risk factors in people with CKD and are often considered the underlying driver of CKD.^2^ For decades, the management of common presentations of CKD consisted of stratifying patient risk by eGFR and UACR and treating with renin-angiotensin system blockade. The lack of additional treatment options diminished the importance of precisely identifying the underlying mechanisms driving an individual patient’s disease process. However, in the past decade multiple CKD therapies have emerged, particularly for those with diabetes and CKD.^1^ With this comes the opportunity to identify which drug classes most benefit individuals. Thus, it is increasingly important to have a more informed appreciation of the underlying structural and molecular disease features at the individual patient level.

While kidney biopsies can provide a definitive tissue-based diagnosis of CKD etiology, in routine clinical practice they are typically only obtained if there is clinical suspicion for a specific diagnosis that could impact management, such as glomerulonephritis. In clinically-indicated kidney biopsy studies of individuals with diabetes, an alternate diagnosis may be present up to 50% of the time.^3^ However, data are lacking regarding the impact of kidney biopsies on diagnosis in people with CKD currently managed by clinical impression only.

The Kidney Precision Medicine Project (KPMP) consortium is obtaining research kidney biopsies from adults with common clinical presentations of CKD.^4^ All biopsies underwent prospective, structured clinico-pathologic adjudication to establish a tissue-based diagnosis of the participant’s CKD, scoring of pathological features, and molecular interrogation, with the aims of identifying novel disease subgroups and discerning underlying disease mechanisms. As most KPMP biopsies were not clinically-indicated, this represents a unique opportunity to garner novel insights into the pathology and molecular mechanisms underlying common CKD presentations.

Here we present findings for 258 KPMP participants with CKD who had undergone clinico-pathological adjudication. With a focus on participants with diabetes and CKD, we demonstrate how clinical, pathological, molecular, and biomarker data can be integrated to establish disease etiology, advance mechanistic understanding, and guide prognostication.

## METHODS

### Study design and population

The KPMP is a cohort study of people with common presentations of CKD and AKI who undergo a standardized kidney biopsy at baseline.^4^ Adults with CKD were recruited from academic centers across the United States. Participants with CKD and type 1 or 2 diabetes (T1D, T2D) were enrolled into the “diabetic kidney disease” (DKD) cohort, and participants with hypertension were enrolled into the “hypertension and chronic kidney disease” (HCKD) cohort. Recruitment site principal investigators had discretion to enroll individuals with diabetes and CKD as HCKD if they felt it was clinically appropriate (for example, when the diagnosis of diabetes had been made just recently before study enrollment and was felt unlikely that diabetes was the main contributor to the participant’s CKD).

### Kidney biopsy and tissue preparation

Kidney biopsies were performed percutaneously by KPMP-certified operators, with up to three biopsy cores obtained.^4^ One diagnostic core was processed locally at the recruitment site under standard clinical protocols using light microscopy (formalin-fixed, paraffin embedded tissue blocks), immunofluorescence (frozen tissue blocks), and electron microscopy (2.5% glutaraldehyde-fixed, epoxy-embedded tissue blocks). Biopsy tissues were evaluated by recruitment site pathologists to establish a clinical diagnosis, which was shared with the study participant via their clinical care team. Digital whole-slide and electron microscopy images of the biopsy slides were used for KPMP clinico-pathological consensus adjudication. Remaining kidney biopsy cores were allocated to KPMP Tissue Interrogation Sites for molecular interrogation, including single-cell RNA sequencing (scRNAseq) and single-nucleus RNA sequencing (snRNAseq).^5,6^

### KPMP biopsy adjudication

Kidney biopsies underwent a formal, prospective, standardized adjudication process to establish a primary clinico-pathological diagnosis.^7^ Information on the participant’s clinical presentation, including blood pressure readings, body mass index, past medical history, medication history, and laboratory values (creatinine-based eGFR, UACR, urine protein creatinine ratio [UPCR], and hemoglobin A1c), were reviewed independently by two KPMP nephrologists. In parallel, the participant’s kidney biopsy slides and recruitment site biopsy report were reviewed independently by two KPMP kidney pathologists, with semiquantitative assessment of glomerular, tubulointerstitial, and vascular features. KPMP nephrologists and pathologists then convened to reach a consensus on the primary cause of the participant’s CKD based on their clinical presentation and pathologic findings from the following diagnostic categories: “diabetic nephropathy”, “hypertension-associated kidney disease”, “other”, or “indeterminate”.^7^

Diabetic nephropathy was defined using Renal Pathology Society criteria.^8^ The diagnosis of hypertension-associated kidney disease was based on a constellation of pathologic features including arterionephrosclerosis, arteriolar hyalinosis and smooth muscle cell hyperplasia, and arterial intimal fibrosis, together with ischemic glomerular (eg., perihilar sclerosis, wrinkling of basement membranes, glomerular tuft retraction) and tubulointerstitial (eg., subcapsular fibrosis) changes.^9^ A primary diagnostic category of “other” was used when biopsy features indicated an alternate primary cause of CKD, while “indeterminate” was selected in cases without predominant classical features of diabetic nephropathy or hypertension-associated kidney, nor clear evidence of alternative kidney pathology.

When KPMP adjudicators reached a different primary diagnosis than the recruitment site pathologist or felt additional clinical evaluation was warranted, the recruitment site principal investigator was informed so that the study participant and their clinical health care professional could also be informed.

### Tubulointerstitial descriptor scoring

Kidney biopsies underwent detailed quantitative assessment of a comprehensive set of 64 tubulointerstitial features independent of the adjudication process, developed based on the NEPTUNE Digital Pathology Scoring System.^10^ Tubulointerstitial descriptor terms and associated definitions were agreed upon by KPMP kidney pathologists by consensus (**Supplemental Data**). Biopsy images were reviewed by two KPMP pathologists, a primary scorer and a quality control scorer, with discrepancies discussed and resolved through discussion and a third pathologist consulted if necessary. Pathologists who completed descriptor scoring did not have access to adjudicated clinico-pathological diagnoses.

### Clinical and laboratory variables

Demographic information and medical history were obtained from participants via KPMP standardized questionnaires completed at the Recruitment Sites. Blood and urine samples were collected at enrollment and laboratory measurements were performed at the Central Laboratory at the University of Washington. Serum creatinine at enrollment was measured using the Beckman Coulter AU5812 automated chemistry analyzer, from which creatinine-based eGFR was calculated using the 2021 CKD-EPI equation.^11^ When a serum sample was not available for creatinine-based eGFR calculation, the most recent eGFR value from the electronic medical record was used (63 participants, 24.4%).

Urine albumin and protein were measured from spot urine samples via immunoturbidimetry and reaction with pyrogallol red and molybdate, respectively. Urine creatinine was measured using the Jaffe reaction with alkaline picrate. From these measures, UACR and UPCR were calculated. When spot urine samples were not available, 24-hour timed urine samples were used (2 participants, 0.8%). When no urine samples were available, the most recent UACR and UPCR values from the electronic medical record were used (62 participants, 24.0%). Plasma concentrations of kidney injury molecule-1 (KIM-1), tumor necrosis factor receptor 1 (TNFR1), and tumor necrosis factor receptor 2 (TNFR2) were measured from enrollment samples using the Mesoscale Diagnostic SQ120 platform (**Supplemental Data**).

### Single-nucleus and single-cell RNA sequencing

Single-cell RNAseq and snRNAseq datasets were generated from kidney tissue samples as described in Lake et al.^5,6^ 50 scRNAseq and 65 snRNAseq samples were included in this study. Differential gene expression analysis was performed on the sc/snRNA data by taking sample-level or cell type-level pseudo-bulk data for each patient. For snRNA data, genes were filtered using the edgeR method implemented in decoupler-py (v1.6) to filter out lowly-expressed genes. Statistical testing was performed using pydeseq2 (v0.4.12) with the assay (10X-multiome or 10XsnRNA) as a covariate. Multiple testing correction is directly done by the package. The Univariate Linear Model method (ULM) implemented in decoupler-py was used for gene set enrichment analyses using the MSigDB hallmarks gene sets, and multiple testing correction was performed using the Benjamin-Hochberg method. DESeq2 R package (v1.50.1) was used for the differential expression analysis of scRNAseq data. Pathway enrichment was performed using decoupleR R package. Genes along with the t-statistic values from DESeq2 output were used as input for decoupleR pathway enrichment analysis using (ULM) model. Pathways enriched from MSigDB hallmarks gene sets (p value < 0.05) were considered as significant.

Correlations of MSigDB pathway activities with sample-level metadata (i.e. pathology descriptor scores or the diabetes signature) were extracted from linear mixed effects models (lme4 v1.1-37 and r2glmm v0.1.2.9001) where the pathway activities were modeled with the sample metadata as fixed effect and the assay as random intercept. Pathway activities in snRNA were computed for each sample pseudobulk with decoupler-py as described above using log1p-transformed normalized counts. This used a subset of samples with both snRNA and biomarker data (n=46).

### Statistical analysis

Descriptive analyses were performed to characterize clinical, laboratory, and pathological features by enrollment cohort and adjudicated diagnosis. Differences in biopsy features by adjudicated clinico-pathological diagnosis were assessed using the chi-square test (or Fisher’s exact test when counts were sparse) for categorical variables, and the Wilcoxon rank-sum test for continuous variables. Two-sided p-values <0.05 were considered statistically significant.

Penalized logistic regression models (LASSO) with 10-fold cross-validation were applied to develop a “diabetic nephropathy signature” distinguishing diabetic nephropathy from other causes of CKD in diabetes in the KPMP. The initial model included age, sex, hemoglobin A1c, eGFR, and log2(UACR), as well as the plasma biomarkers log2(KIM-1), log2(TNFR1), and log2(TNFR2), from which a subset of variables was selected using LASSO as best classifying diabetic nephropathy. Plasma KIM-1, TNFR1, and TNFR2 were included as these biomarkers have been approved by the Food and Drug Administration for assessment of risk of progressive kidney function decline as part of the KidneyIntelX.dkd test.^12,13^ Receiver operating characteristic curve and area under the curve analyses were used to evaluate the model’s discriminatory ability in the KPMP.

The resulting model was then applied to participants from the Chronic Renal Insufficiency Cohort (CRIC) with diabetes and available biomarker measurements (n=229), generating each participant’s predicted probability of having diabetic nephropathy. Cox proportional hazard models were used to determine if the predicted probability of having diabetic nephropathy was associated with time to the composite outcome of dialysis, transplantation, or 50% eGFR decline from baseline, adjusting for demographic and clinical variables: age, sex, race, ethnicity, hypertension history, cardiovascular disease history, smoking history, body mass index, systolic blood pressure, renin-angiotensin blockade use, anti-diabetes medication use, hemoglobin A1c, eGFR, UACR.^14,15^ Bootstrapped C-indices were then used to assess model discrimination and compare performance of clinical variables alone versus clinical variables combined with the diabetic nephropathy signature in predicting the composite kidney outcome.

## RESULTS

### Participant characteristics

Clinico-pathological adjudication was completed for 258 KPMP CKD participants (**Table 1**). Participants had a mean age of 61 years, 134 (52%) were male; 118 (46%) self-identified as White, 89 (35%) as Black or African American, and 66 (26%) as Latino. Overall, at enrollment, participants had mean eGFR 54 mL/min/1.73m^2^ and 181 (70%) had mean eGFR <60 mL/min/1.73m^2^. Participants enrolled as DKD (n=183) had mean eGFR 56 mL/min/1.73m^2^ and median UACR 432 mg/g; 135 (74%) had UACR ≥30 mg/g. Participants enrolled as HCKD (n=75) had mean eGFR 52 mL/min/1.73m^2^ and median UACR 30mg/g; 37 (49%) had UACR ≥30 mg/g.

**Table 1.** Baseline characteristics of KPMP participants with CKD. Baseline characteristics of N=258 participants for whom adjudication has been completed, by enrollment cohort.

|  | **Overall**<br>**(n=258)** | **DKD**<br>**(n=183)** | **HCKD**<br>**(n=75)** |
| --- | --- | --- | --- |
| **Male, N (%)** | 134 (51.9) | 97 (53.0) | 37 (49.3) |
| **Age, years, mean (SD)** | 60.5 (11.8) | 59.5 (12.1) | 62.7 (10.9) |
| **Race, N (%)** |  |  |  |
| **Asian** | 9 (3.5) | 7 (3.8) | 2 (2.7) |
| **Black or African-American** | 89 (34.5) | 57 (31.1) | 32 (42.7) |
| **White** | 118 (45.7) | 86 (47.0) | 32 (42.7) |
| **Other/Don't Know** | 39 (15.1) | 30 (16.4) | 9 (12.0) |
| **Latino, N (%)** | 66 (25.7) | 56 (30.8) | 10 (13.3) |
| **BMI, kg/m<sup>2</sup>, mean (SD)** | 31.1 (6.7) | 31.2 (6.8) | 30.8 (6.3) |
| **Diabetes history, N (%)** | 193 (75.1) | 178 (97.8) | 15 (20.0) |
| **Diabetes type among people with diabetes, N (%)** |  |  |  |
| **Type 1** | 17 (8.8) | 16 (9.0) | 1 (6.7) |
| **Type 2** | 176 (91.2) | 162 (91.0) | 14 (93.3) |
| **Diabetes duration, years, mean (SD)** | 18.0 (11.5) | 18.7 (11.1) | 9.7 (12.5) |
| **Hypertension history, N (%)** | 241 (93.8) | 166 (91.2) | 75 (100.0) |
| **Hypertension duration, years, mean (SD)** | 17.1 (13.0) | 15.4 (12.4) | 20.4 (13.6) |
| **Diabetic retinopathy history, N (%)** | 84 (43.5) | 83 (46.6) | 1 (6.7) |
| **Diabetic neuropathy history, N (%)** | 81 (42.0) | 77 (43.3) | 4 (26.7) |
| **Peripheral vascular disease history, N (%)** | 20 (10.4) | 19 (10.7) | 1 (6.7) |
| **Coronary artery disease history, N (%)** | 22 (8.6) | 15 (8.2) | 7 (9.3) |
| **High cholesterol history, N (%)** | 163 (63.7) | 117 (64.6) | 46 (61.3) |
| **Heart failure history, N (%)** | 17 (6.6) | 10 (5.5) | 7 (9.3) |
| **Stroke history, N (%)** | 11 (4.3) | 7 (3.8) | 4 (5.3) |
| **Diuretic use, N (%)** | 110 (42.6) | 82 (44.8) | 28 (37.3) |
| **ACEi/ARB use, N (%)** | 199 (77.1) | 146 (79.8) | 53 (70.7) |
| **SGLT2i use, N (%)** | 100 (38.8) | 91 (49.7) | 9 (12.0) |
| **Systolic blood pressure, mmHg, mean (SD)** | 137.15 (12.71) | 137.49 (12.68) | 136.33 (12.82) |
| **Diastolic blood pressure, mmHg, mean (SD)** | 75.67 (9.84) | 75.79 (9.44) | 75.37 (10.82) |
| **Hemoglobin A1c, %, mean (SD)** | 7.33 (1.81) | 7.78 (1.80) | 5.84 (0.71) |
| **eGFR, mL/min/1.73m<sup>2</sup>, mean (SD)** | 54 (22) | 56 (24) | 52 (16) |
| **eGFR category, N (%)** |  |  |  |
| **eGFR 15-29** | 12 (4.7) | 6 (3.3) | 6 (8.0) |
| **eGFR 30-44** | 88 (34.1) | 72 (39.3) | 16 (21.3) |
| **eGFR 45-59** | 81 (31.4) | 51 (27.9) | 30 (40.0) |
| **eGFR > 60** | 77 (29.8) | 54 (29.5) | 23 (30.7) |
| **Albuminuria category, N (%)** |  |  |  |
| **< 30 mg/g Cr** | 77 (30.9) | 40 (22.9) | 37 (50.0) |
| **30-299 mg/g Cr** | 65 (26.1) | 43 (24.6) | 22 (29.7) |
| **≥ 300 mg/g Cr** | 107 (43.0) | 92 (52.6) | 15 (20.3) |
| **UACR mg/g Cr, median [IQR]** | 198 [16, 927] | 432 [41, 1508] | 30 [6, 215] |
| **UPCR mg/g Cr, median [IQR]** | 418 [123, 1321] | 694 [170, 2235] | 169 [74, 453] |

Don't know/declined to answer, N (%): race 3 (1.2); Latino 5 (); diabetes history 2 (0.8); hypertension history 2 (0.8); diabetic retinopathy history 8 (4.1); diabetic neuropathy history 8 (4.1); peripheral vascular disease history 8 (4.1); coronary artery disease history 6 (2.3); high cholesterol history 12 (4.7); heart failure history 8 (3.1); stroke history 3 (1.2)
Missingness N (%): Latino 1 (0.4); BMI 3 (1.2); diabetes history 1 (0.4); diabetes duration 13 (6.7); hypertension history 1 (0.4); hypertension duration 57 (22.1); coronary artery disease history 1 (0.4); stroke history 1 (0.4); high cholesterol history 2 (0.8); heart failure history 1 (0.4); hemoglobin A1c 30 (11.6); albuminuria category 9 (3.5); UACR 9 (3.5); UPCR 20 (7.8)

### Kidney biopsy adjudication results

Among the 183 participants enrolled as DKD, 102 (56%) had a primary adjudicated diagnosis of diabetic nephropathy, 35 (19%) had a primary adjudicated diagnosis of hypertension-associated kidney disease, 18 (10%) had other adjudicated categories, and 28 (15%) had indeterminate findings (**Figure 1A**). Among the 75 participants enrolled as HCKD, 42 (56%) had a primary diagnosis of hypertension-associated kidney disease, 3 (4%) had a primary diagnosis of diabetic nephropathy, 12 (16%) had other diagnoses, and 18 (24%) had indeterminate findings. For participants adjudicated as “other”, underlying etiologies included focal segmental glomerulosclerosis, IgA nephropathy, membranous nephropathy, and fibrillary glomerulonephritis, among others (**Figure 1B**). Recruitment site principal investigators had discretion to enroll individuals with diabetes and CKD as HCKD if they felt it was clinically appropriate (for example, when the diagnosis of diabetes had been made just recently before study enrollment and was felt unlikely that diabetes was the main contributor to the participant’s CKD).

**Figure 1.**
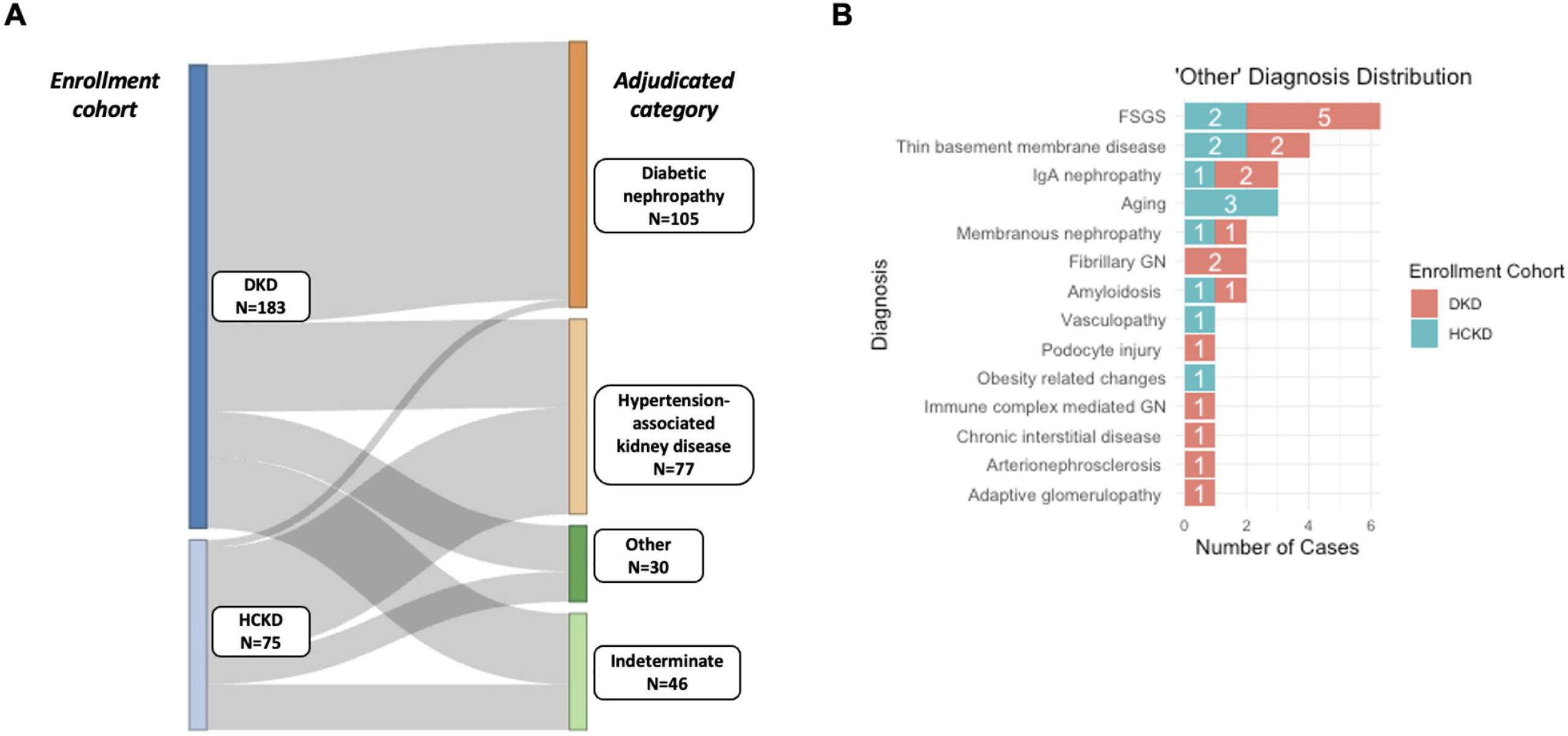
Adjudicated biopsy diagnoses by enrollment cohort in the KPMP. **(A)** Sankey plot of KPMP participants showing enrollment cohort and final adjudicated diagnosis. **(B)** Summary of “other” diagnoses identified in KPMP participants. DKD: diabetic kidney disease; HCKD: hypertension and chronic kidney disease

As a sensitivity analysis, we examined the distribution of adjudicated categories by participants’ self-reported diabetes status (**Supplemental Figure 1**). Among the 193 participants who reported having diabetes, 103 (53%) had a primary diagnosis of diabetic nephropathy, 42 (22%) had hypertension-associated kidney disease, 19 (10%) had other diagnoses, and 29 (15%) had indeterminate findings. Among the 62 participants who reported not having diabetes, 34 (62%) had hypertension-associated kidney disease, 1 (2%) had diabetic nephropathy, 11 (18%) had other diagnoses, and 16 (26%) had indeterminate findings. Overall, results were similar when data were analyzed by enrollment cohort.

### Kidney biopsy pathologic findings

A broad range of glomerular, tubulointerstitial, and vascular findings was observed across KPMP CKD biopsies (**Figure 2**). Among the set of standard clinically-assessed biopsy features, participants with diabetic nephropathy also had significantly greater segmental sclerosis/collapse (present in 39% vs 21%) and more severe tubulointerstitial fibrosis (>1+ in 64% vs 37%), tubular atrophy (>1+ in 63% vs 36%), and arteriolar hyalinosis (severe in 53% vs 29%) compared to those with hypertension-associated kidney disease (p-value <0.01 for all; **Figure 2A**).

**Figure 2.**
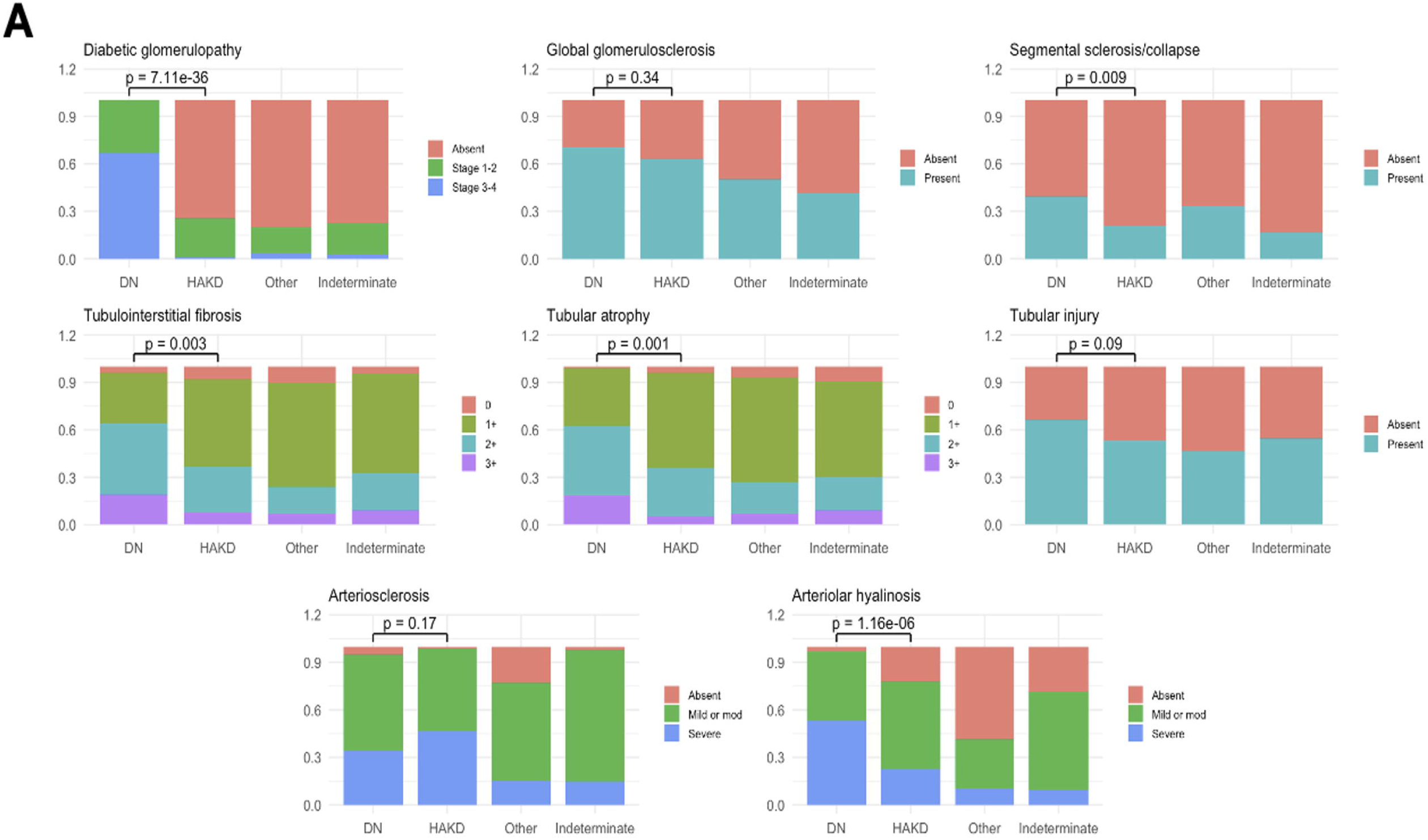
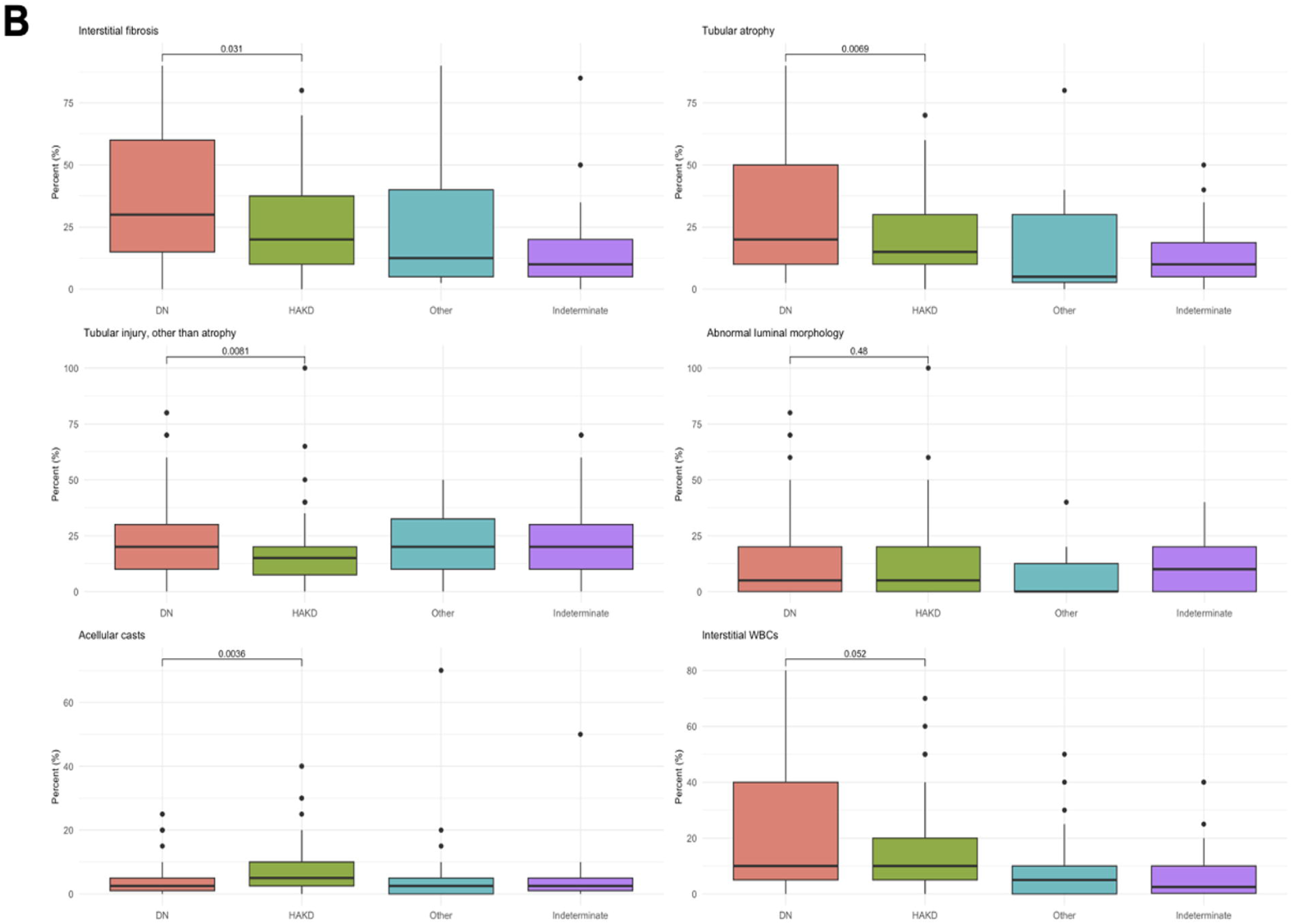
Biopsy features of KPMP participants with CKD by adjudicated diagnosis. **(A)** Distribution of standard clinically-assessed glomerular, tubulointerstitial, and vascular biopsy features by adjudicated diagnosis. **(B)** Distribution of select quantitative tubulointerstitial descriptors by adjudicated diagnosis. Differences in categorical biopsy features between participants with adjudicated diabetic nephropathy and hypertension-associated kidney disease were assessed using the Fischer’s exact test and differences in continuous features were assessed using the Wilcoxon-rank sum test.

Tubulointerstitial features underwent further detailed assessment with comprehensive, quantitative scoring (**Figure 2B, Supplemental Figure 2**). Assessing tubulointerstitial descriptors, participants with diabetic nephropathy had significantly greater interstitial fibrosis (mean 35% [SD 25%] vs 26% [20%] of interstitium), tubular atrophy (31% [24%] vs 21% [17%] of interstitium), and tubular injury other than atrophy (25% [19%] vs 18% [17%] of interstitium) than those with hypertension-associated kidney disease (p-value <0.05 for all, **Figure 2B, Supplemental Figure 2**). There was also greater white blood cell interstitial infiltration among participants with diabetic nephropathy, though this difference approached but did not reach statistical significance (23% [23%] vs 15% [16%] of interstitium; p-value = 0.052). Additionally, acellular casts were more prevalent in hypertension-associated kidney disease than diabetic nephropathy (9% [9%] vs 5% [6%] of cortex; p-value = 0.004).

Interstitial edema and tubular simplification were more often present in diabetic nephropathy, and thyroidization-type tubular atrophy was more often present in hypertension-associated kidney disease (p-value <0.05 for all, **Supplemental Figure 2**).

### Development of a clinical and biomarker signature of diabetic nephropathy

For KPMP participants with available biomarker data, a parsimonious signature distinguishing diabetic nephropathy (n=76) from other causes of CKD in diabetes (n=60) was developed using LASSO with 10-fold cross-validation (**Supplemental Table 1**). The variables age, sex, HbA1c, eGFR, UACR, KIM-1, TNFR1, and TNFR2 were initially provided to the model. The model ultimately selected for age, sex, HbA1c, UACR, KIM-1, and TNFR1, and yielded an AUC of 0.82 (95% CI 0.75-0.89), consistent with good discrimination of diabetic nephropathy (**Figures 3A,B**).

**Figure 3.**
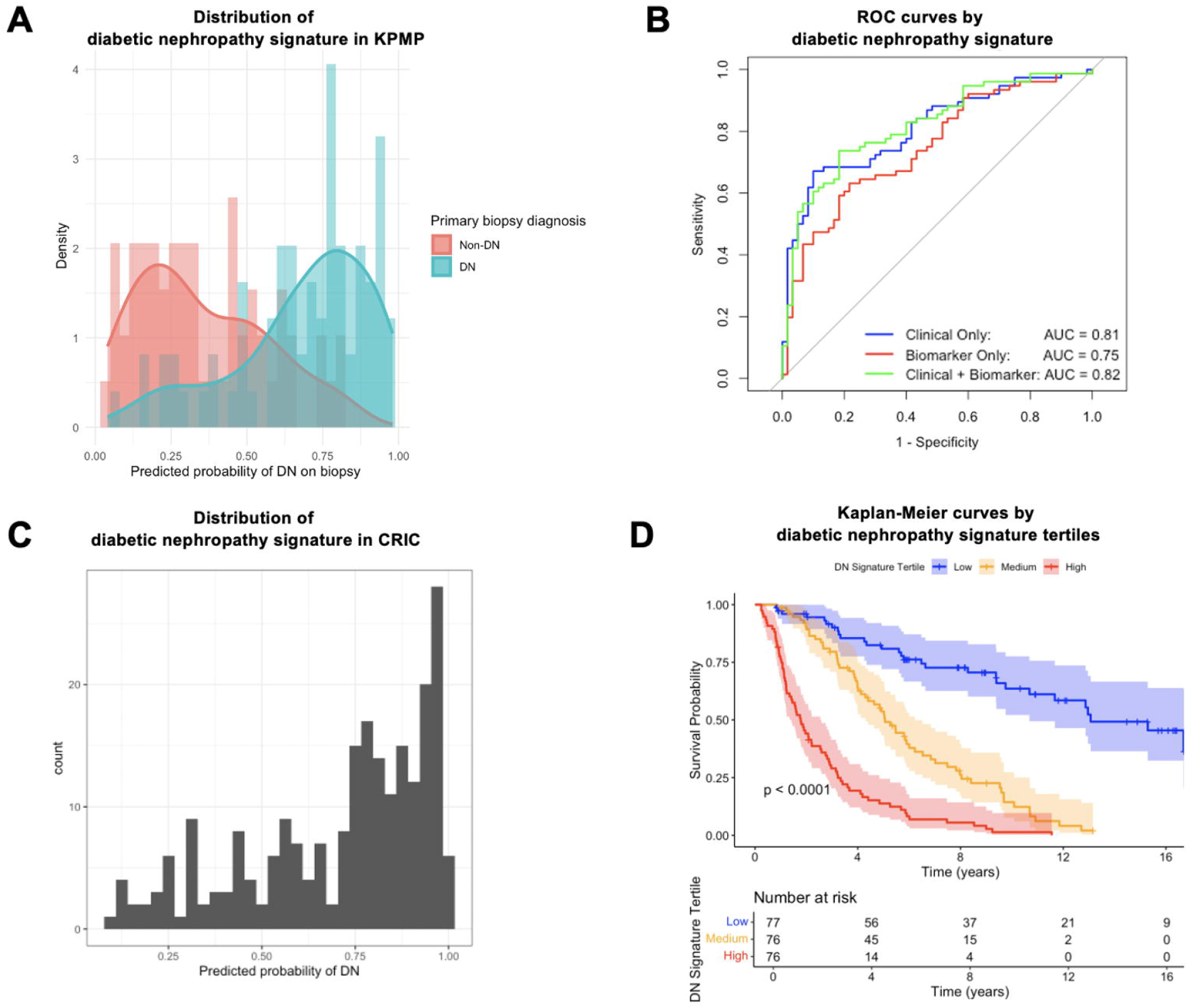
Development of a clinical and biomarker signature reflecting predicted probability of DKD. **(A)** Distribution of the “diabetic nephropathy signature” reflecting predicted probability of diabetic nephropathy in KPMP participants with diabetes and CKD, with and without biopsy-proven diabetic nephropathy (n=136). **(B)** Receiver operating characteristic curve showing discriminatory ability of the diabetic nephropathy signature in the KPMP. **(C)** Distribution of the diabetic nephropathy signature in CRIC. **(D)** Kaplan-Meier curves comparing rates of incident dialysis, transplantation, or ≥50% eGFR decline by diabetic nephropathy signature tertile in CRIC. No USRDS data is used.

CRIC participants (n=229) had a mean age of 58 years, 153 (67%) were male; 63 (28%) identified as non-Hispanic White, 77 (34%) identified as non-Hispanic Black, and 85 (37%) identified as Hispanic (**Supplemental Table 2**). At baseline, participants had mean HbA1c 7.4%, mean eGFR 39 mL/min/1.73m^2^, median UACR 446 mg/g. The diabetic nephropathy signature was applied to CRIC participants, then examined for associations with incident dialysis, transplantation, or eGFR decline ≥50% (**Figure 3C**) In total, 165 (72%) CRIC participants experienced the outcome over a period of 17.1 years (median 4.0 years). The diabetic nephropathy signature was significantly associated with the composite kidney outcome (HR 1.48 [95% CI 1.27-1.73] per 10% higher predicted probability of diabetic nephropathy), adjusting for demographic and clinical variables; significant differences by tertile of diabetic nephropathy signature were also apparent (**Figure 3D**). The model incorporating clinical variables combined with the diabetic nephropathy signature better predicted the composite kidney outcome than the model using clinical variables alone (C-index 0.793 vs 0.778; delta C-index 0.0147 [95% CI 0.0003, 0.0290]).

### Identification of molecular pathways in diabetic nephropathy

Kidney single-cell (sc) and single-nucleus (sn) RNA sequencing data from KPMP CKD biopsies with diabetic nephropathy (sn=29, sc=19) were compared to biopsies adjudicated as hypertension-associated kidney disease (sn=26, sc=17) or indeterminate (sn=10, sc=14) to identify molecular pathways that characterize diabetic nephropathy (**Figure 4A, Supplemental Figure 3**). Epithelial mesenchymal transition, MTORC1 signaling, and inflammatory response pathways were generally upregulated in diabetic nephropathy across multiple cell types. Oxidative phosphorylation and fatty acid metabolism showed a nephron segment specific regulation, with repression in proximal and activation in distal tubular segments.

**Figure 4.**
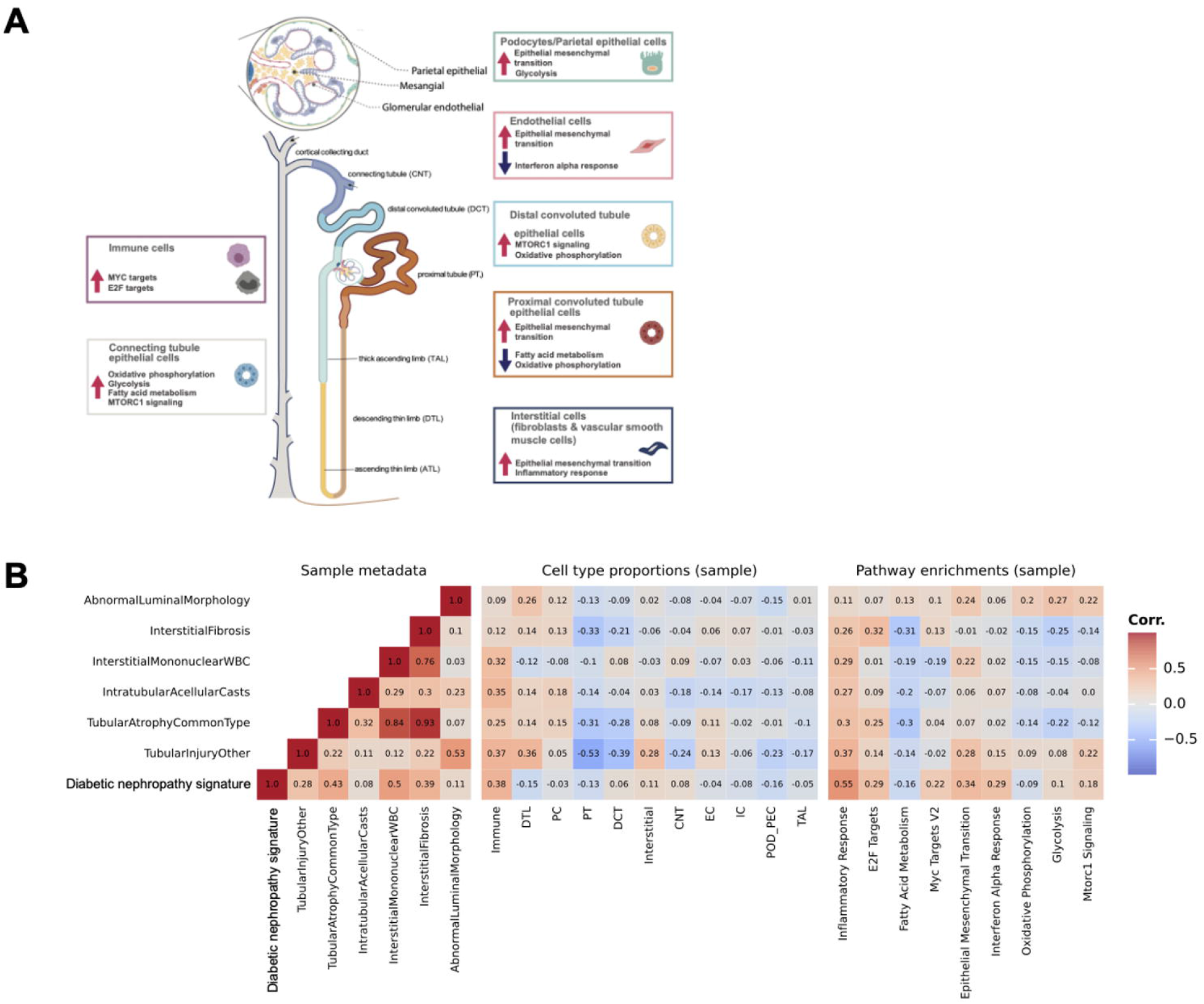
Transcriptomic data analyses in diabetic nephropathy. **(A)** Top differentially-expressed transcriptomic pathways in KPMP participants with CKD and diabetic nephropathy compared to those with hypertension-associated kidney disease or indeterminate diagnoses. Pathways shown were differentially expressed in both single-cell and single-nucleus RNA sequencing data. Upregulated pathways in diabetic nephropathy are depicted with a red arrow whereas downregulated pathways are depicted with a blue arrow. **(B)** Correlations of diabetic nephropathy signature with kidney cell types, tubulointerstitial biopsy features, and diabetic nephropathy-enriched transcriptomic pathways.

We then applied the diabetic nephropathy signature to KPMP biopsies with single-nucleus RNA sequencing data and examined correlations between these, as well as of quantitatively-assessed tubulointerstitial biopsy features (**Figure 4B**). Overall, positive correlations were observed between the diabetic nephropathy signature and tubulointerstitial features, with the strongest correlations observed with white blood cell interstitial infiltration (*r*=0.5), tubular atrophy (*r*=0.43), and interstitial fibrosis (*r*=0.39). Among kidney cell types, the diabetic nephropathy signature correlated most strongly with higher proportion of immune cells (*r*=0.38). Additionally, the diabetic nephropathy signature exhibited correlations with molecular pathways differentially expressed in diabetic nephropathy. The strongest correlations were observed between the diabetic nephropathy signature and inflammatory response (*r*=0.55), epithelial mesenchymal transition (*r*=0.34), interferon alpha response (*r*=0.29), and E2F targets (*r*=0.29) pathways. While generally weaker correlations were observed between individual tubulointerstitial features and molecular pathways, the strongest of these were observed with inflammatory response (*r*=0.11-0.37). Furthermore, the diabetic nephropathy signature correlated with altered proximal tubular cells reflecting tubular injury, specifically adaptive (*r*=0.35) and failed repair states (*r*=0.41); tubulointerstitial features also correlated with tubular cell states (**Supplemental Figure 4**).

## DISCUSSION

In the KPMP, kidney biopsies performed in adults with common clinical presentations of CKD more precisely defined the underlying structural and molecular features of the disease process by establishing a tissue-based diagnosis. The adjudication process produced categories that were meaningful at the histologic and molecular level. Analyses of kidney pathological features and transcriptional data revealed greater severity of tubulointerstitial lesions and upregulation of inflammatory pathways in diabetic nephropathy compared to other etiologies of CKD. The probability of diabetic nephropathy could be captured by a non-invasive signature which was associated with adverse long-term kidney outcomes in an independent cohort of adults with diabetes and CKD. Together, these findings demonstrate that careful characterization of patients with common presentations of CKD can reveal differences in kidney structure and function that have important prognostic and therapeutic implications.

Biopsies performed in KPMP participants with CKD often yielded diagnoses that differed from clinicians’ expectations. Among participants enrolled as DKD, only approximately half had a primary biopsy diagnosis of diabetic nephropathy, with hypertension-associated kidney disease as the next most common diagnosis. This observation is consistent with prior studies of clinical biopsies reporting a similar prevalence of diabetic nephropathy, as well as with studies describing a vascular pathological subphenotype of diabetes and CKD consisting of primary hypertensive and ischemic changes.^3,16^ Similarly among participants enrolled as HCKD, half had a primary diagnosis of hypertension-associated kidney disease. Additionally, both participants enrolled as DKD and HCKD often had indeterminate findings on biopsy which precluded establishment of a primary diagnosis. These cases illustrate the limitations of traditional biopsy-based diagnostic frameworks for common CKD presentations in which the dramatic clinical changes that typically prompt a biopsy (such as rapidly rising albuminuria, declining eGFR, or hematuria) may not be present.^7^ Moreover, kidney biopsy regularly identified unexpected diagnoses–such as IgA nephropathy, membranous nephropathy, and fibrillary glomerulonephritis–for which alternative treatment approaches are indicated.

While there was overlap in pathological patterns observed across CKD biopsies, diabetic nephropathy generally exhibited more severe tubulointerstitial lesions than hypertension-associated kidney disease. Evaluation of independently-scored pathological features showed that participants with diabetic nephropathy had increased interstitial fibrosis, tubular atrophy, and tubular injury other than atrophy, as well as a trend towards greater interstitial inflammatory infiltrate. These findings have been associated with CKD progression.^17,18^ Moreover, participants with diabetic nephropathy were more likely to have segmental glomerulosclerosis and more severe arteriolar hyalinosis, which have also been associated with poor outcomes.^19–21^

Overall, these observations highlight how diabetic nephropathy, despite being primarily being defined by classic diabetic glomerular lesions, also presents with distinctive and clinically-meaningful tubulointerstitial findings.^7^ This has also been previously documented in research kidney biopsy studies among participants with type 1 diabetes, where even at earlier stages, thickening of glomerular basement membrane parallel that of the tubular basement membrane.^22^ Our work also sheds light on the histopathology of hypertension-associated kidney disease, which remains a relatively unexplored area despite the high population prevalence.^23^ In addition to reporting on standard glomerular, tubulointerstitial, and vascular lesions in hypertension-associated kidney disease, we also observed more intratubular acellular casts, microcystic lesions, and thyroidization type tubular atrophy, which to our knowledge have not previously been described.

An important finding is that a combination of clinical features and plasma biomarkers was sufficient to develop a minimally-invasive signature that distinguished diabetic nephropathy from other causes of CKD in diabetes in the KPMP. This signature contained information that health care professionals commonly use in clinical practice to ascertain whether diabetes is contributing to a patient’s CKD, including albuminuria and HbA1c, as well as two plasma biomarkers, KIM-1 and TNFR1, that reflect underlying histologic and molecular features of diabetic nephropathy and are associated with kidney function decline in long-term cohort studies and clinical trials.^24–29^ KIM-1 is widely considered a marker of tubular damage and associates with various tubulointerstitial features associated with poor prognosis.^30,31^ TNFR1, a receptor for TNFα, activates immune pathways and is associated with glomerular features and CKD progression in diabetes cohorts.^32–34^ Moreover, predicted probability of diabetic nephropathy was associated with long-term adverse kidney outcomes in CRIC, underscoring the importance of establishing this diagnosis for guiding clinical prognostication and timely treatment initiation. Critically, early KIM-1 and TNFR1 level reduction in response to SGLT2 inhibition in CANVAS were independently associated with better long-term outcomes.^29^ Currently, the treatment of patients with diabetes who have CKD has a “one-size-fits-all” approach–distinguishing risk profiles based on clinico-pathological diagnoses may allow for more personalized therapeutic approaches with greater clinical efficacy.

Analyses of kidney transcriptional data in the KPMP identified consistent enrichment of inflammatory pathways along nephron segments in diabetic nephropathy. Inflammation has long been recognized as playing a key role in the pathogenesis of diabetic nephropathy, with specifically, TNF signaling and interferon gamma signaling identified as important mechanistic drivers.^35^ These results align with our pathologic findings of white blood cell interstitial infiltration, as well as our observations that KIM-1 and sTNFR1 help distinguish diabetic nephropathy from other forms of CKD. Oxidative phosphorylation showed more cell type-specific regulation with repression in proximal but activation in distal nephron segments, consistent with proximal tubular mitochondrial dysfunction as a well-established feature of diabetic nephropathy.^36–38^ Additionally, we observed mild to moderate correlations between the diabetic nephropathy signature, tubulointerstitial biopsy features, and molecular pathways, underscoring links between clinical, biomarker, structural, and functional aspects of diabetic nephropathy. Notably, the strongest correlations were observed between the diabetic nephropathy signature, interstitial white blood cell infiltration on pathology, and molecular inflammatory response and epithelial mesenchymal transition pathways, highlighting the central role of inflammation in diabetic nephropathy.

Overall, our findings suggest that kidney biopsies in patients with CKD provide highly valuable information relevant to diagnosis, prognosis, and disease-driving mechanisms. Biopsy data can have important implications for clinical care relevant in both clinical and research settings. Clinically, the examination of kidney biopsy tissues can refine the underlying driver of an individual’s CKD and give prognostic insights, in some cases identifying unexpected diagnoses who would benefit from individualized clinical therapies.^39–42^ We demonstrate that diabetic nephropathy is characterized by distinct pathological and molecular patterns, and may be associated with worse long-term kidney outcomes. These findings have the potential to inform individualized therapeutic approaches in diabetes and CKD. In the research setting, insight into kidney tissue pathology–directly ascertained by kidney biopsy or predicted by surrogate biomarker signatures–could inform participant selection, treatment allocation, and evaluation of outcomes in CKD studies, including clinical trials of novel therapeutic agents. By identifying the specific underlying cause of CKD, study participants could be better risk-stratified, or selected for targeted therapies. For example, patients with diabetic nephropathy were more likely to have derangements in mTORC signaling, which is a pathway targeted by SGLT2 inhibitors.

KPMP has several strengths, including that it is a multi-center study with diverse participant population with careful adherence to data quality and tissue processing. However, there are limitations, including that some of our participants were recruited after agreeing to a clinically indicated biopsy. Additionally, kidney biopsies were adjudicated off of one core of tissue, which is not standard clinical practice and may have resulted in less biopsy tissue to be reviewed. As KPMP is actively recruiting, long-term outcome data are not yet available for participants, although we were able to identify a cross-sectional signature of the diabetic nephropathy that was associated with increased risk of kidney disease progression in CRIC, thus providing external validation.

In conclusion, kidney biopsies in KPMP participants with CKD yielded clinico-pathological diagnoses that provided meaningful pathologic structural and molecular information. Integration of clinical, pathological, and biomarker data in the KPMP enabled development of a model that predicted diabetic nephropathy and was associated with adverse kidney outcomes in an independent cohort. Overall, these results suggest that kidney biopsies in common clinical presentations of CKD can provide valuable diagnostic, mechanistic, and prognostic information.

## Supporting information

KPMP Collaborator List for Indexing with Submission

## Data Availability

Clinical and histopathological data used in the study are available upon request to the KPMP via kpmp.org. Biomarker and molecular data used in the study are available online at kpmp.org.

https://www.kpmp.org/

## DISCLOSURES

The authors have no relevant conflicts to disclose.

## ACKNOWLEDGMENTS

The study was approved by the University of Washington Institutional Review Board (IRB 20190213). Written informed consent was received from all participants prior to study participation.

The authors acknowledge the University of Michigan Medical School Central Biorepository (RRID:SCR_026845) for providing biospecimen storage, management, and distribution services in support of the research reported in this publication.

A portion of the data reported here have been supplied by the United States Renal Data System (USRDS). The interpretation and reporting of these data are the responsibility of the author(s) and in no way should be seen as an official policy or interpretation of the U.S. government.

## FUNDING

The Kidney Precision Medicine Project (KPMP) is supported by the National Institute of Diabetes and Digestive and Kidney Diseases (NIDDK) through the following grants: U01DK133081, U01DK133091, U01DK133092, U01DK133093, U01DK133095, U01DK133097, U01DK114866, U01DK114908, U01DK133090, U01DK133113, U01DK133766, U01DK133768, U01DK114907, U01DK114920, U01DK114923, U01DK114933, U24DK114886, UH3DK114926, UH3DK114861, UH3DK114915, and UH3DK114937.

Funding for the CRIC Study was obtained under a cooperative agreement from National Institute of Diabetes and Digestive and Kidney Diseases (U01DK060990, U01DK060984, U01DK061022, U01DK061021, U01DK061028,U01DK060980, U01DK060963, U01DK060902 and U24DK060990). In addition, this work was supported in part by: the Perelman School of Medicine at the University of Pennsylvania Clinical and Translational Science Award NIH/NCATS UL1TR000003, Johns Hopkins University UL1 TR-000424, University of Maryland GCRC M01 RR-16500, Clinical and Translational Science Collaborative of Cleveland, UL1TR000439 from the National Center for Advancing Translational Sciences (NCATS) component of the National Institutes of Health and NIH roadmap for Medical Research, Michigan Institute for Clinical and Health Research (MICHR) UL1TR000433, University of Illinois at Chicago CTSA UL1RR029879, Tulane COBRE for Clinical and Translational Research in Cardiometabolic Diseases P20 GM109036, Kaiser Permanente NIH/NCRR UCSF-CTSI UL1 RR-024131, Department of Internal Medicine, University of New Mexico School of Medicine Albuquerque, NM R01DK119199.

We gratefully acknowledge the essential contributions of our patient participants and the support of the American public through their tax dollars. The content is solely the responsibility of the authors and does not necessarily represent the official views of the National Institutes of Health.

## Notes

### Competing Interest Statement

The authors have declared no competing interest.

### Clinical Protocols

https://drive.google.com/file/d/1pFRcWRXYeswjruoG-EBz49zCA9DJPiwu/view

https://drive.google.com/file/d/1bcM_Z0GDLRTLZKXQmDYvAyCm-1tu9zBG/view

