## Supplemental Tables and Figures for "Redefining kidney disease: Clinico-pathological and molecular findings from the Kidney Precision Medicine Project"

**Supplemental Figure 1. Adjudicated biopsy diagnoses by diabetes and CKD status in the KPMP.** Sankey plot of KPMP participants showing diabetes and CKD status and final adjudicated diagnosis.

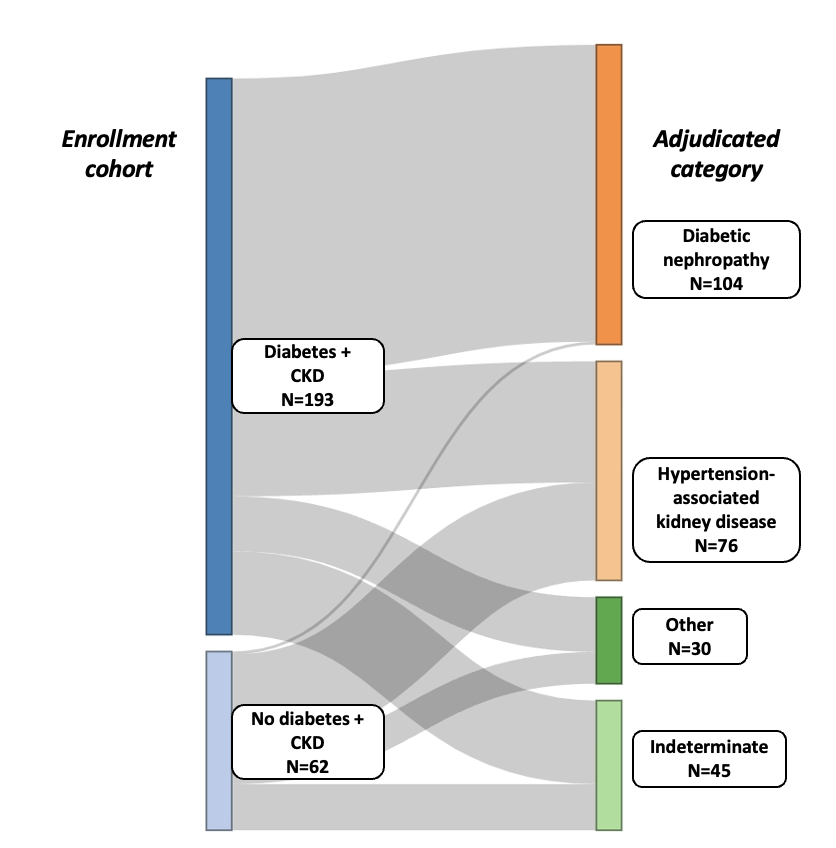

**Supplemental Figure 2. Tubulointerstitial biopsy features of KPMP participants with CKD by adjudicated diagnosis.** Differences in categorical biopsy features between participants with adjudicated diabetic nephropathy and hypertension-associated kidney disease were assessed using the Fischer’s exact test.

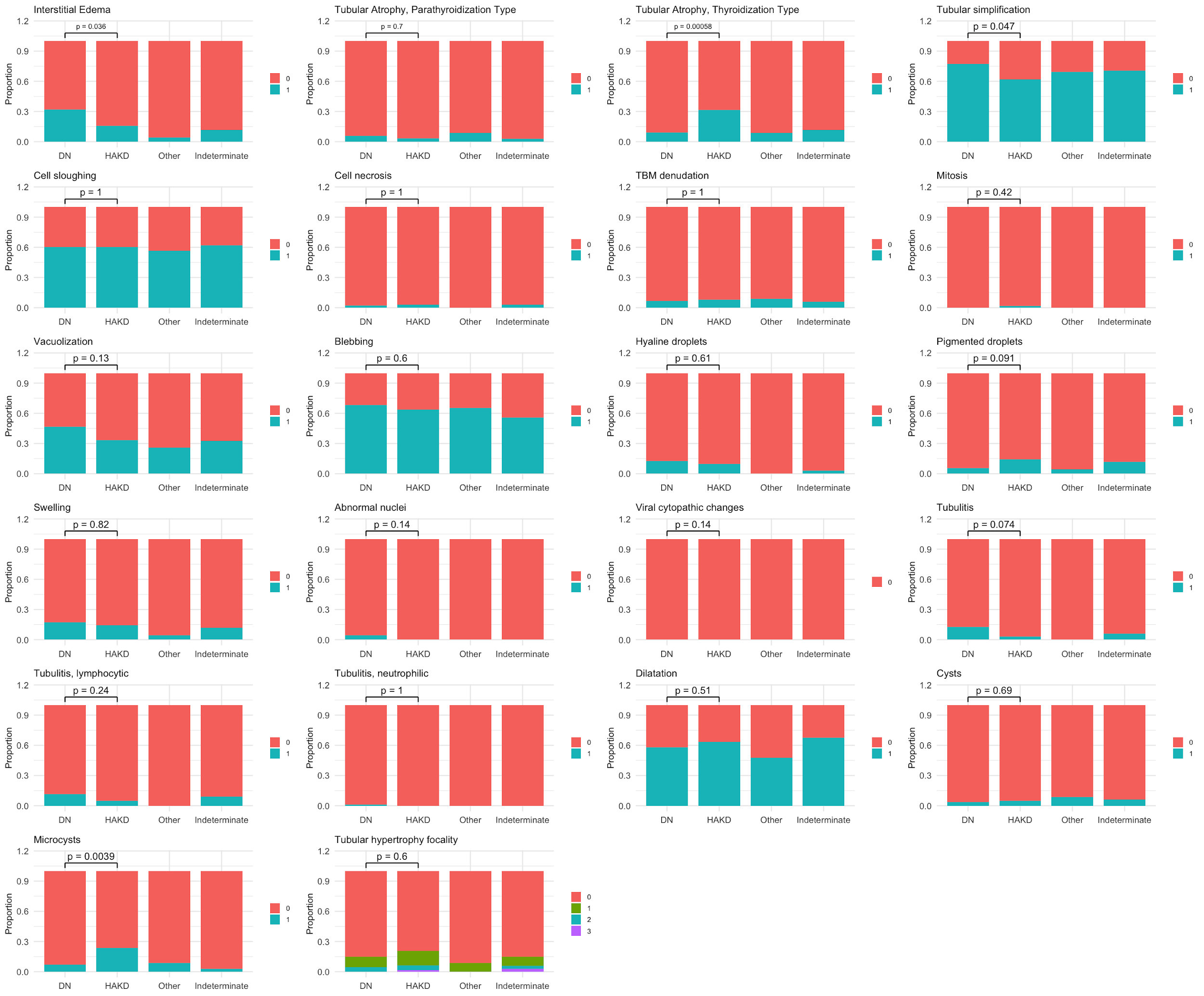

**Supplemental Figure 3. Top differentially-expressed pathways using single-cell and single-nucleus RNA sequencing data comparing participants with diabetic nephropathy to those with hypertension-associated kidney disease or indeterminate diagnoses.**

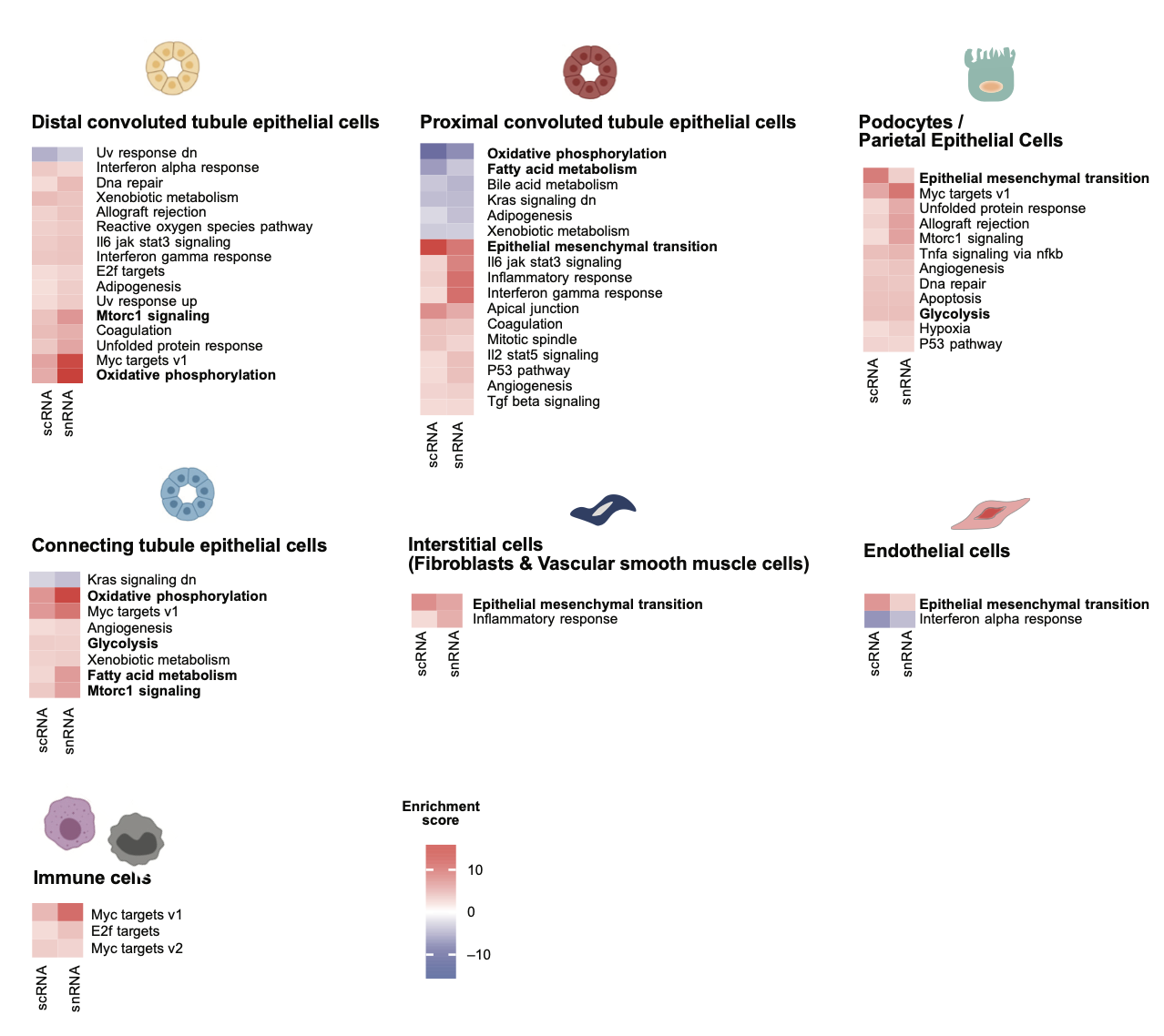

**Supplemental Figure 4. Correlations of diabetic nephropathy signature with proximal tubular cell states, tubulointerstitial biopsy features, diabetic nephropathy-enriched proximal tubular transcriptomic pathways.** aPT: adaptive proximal tubular cell; frPT: failed repair proximal tubular cell; PT-S3: proximal tubule segment S3; PT-S2: proximal tubule segment S2; dPT: degenerative proximal tubular cell; PT-S1: proximal tubule segment S1; cycPT: cycling proximal tubular cell

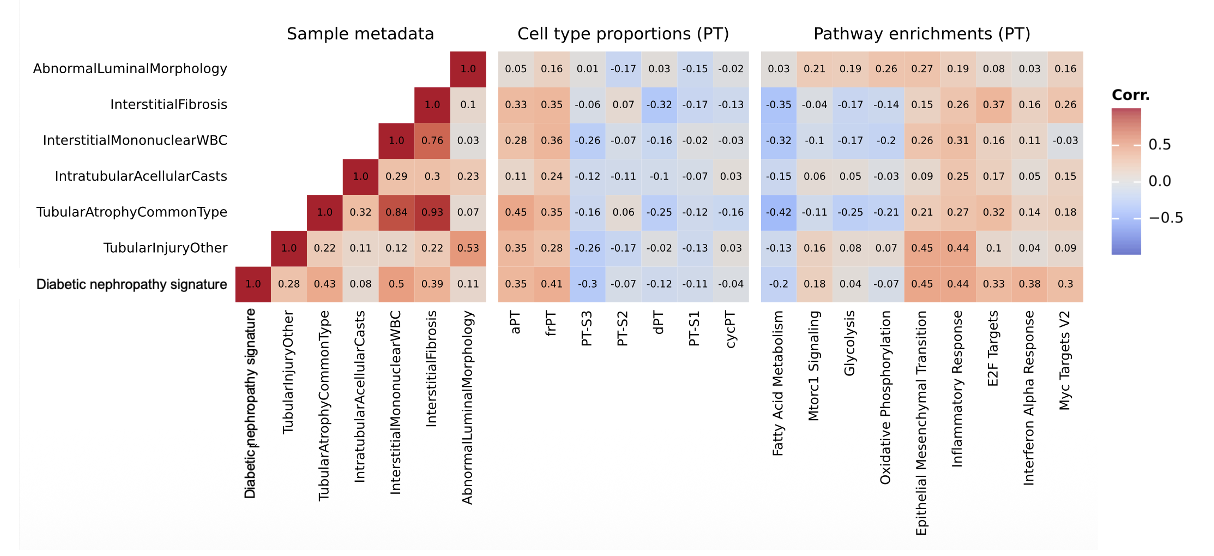
